# Biomechanical assessment of upright birthing positions using marker-based and markerless motion capture systems

**DOI:** 10.1101/2025.02.18.25322290

**Authors:** Lauren Haworth, Inderjeet Kaur, Maimoona Ahmed, Anastasia Topalidou

## Abstract

Understanding the effects of birthing positions on labour is crucial for optimising maternal and foetal outcomes. Upright positions are encouraged but their biomechanics are not fully understood. Biomechanical changes during labour can make certain positions more or less favourable depending on individual physical characteristics. Understanding these factors is essential for tailoring strategies that enhance maternal comfort and facilitate labour. This study aimed to quantify the biomechanics of seven common upright birthing positions, comparing their biomechanical characteristics and evaluating the sensitivity and accuracy of marker-based and markerless motion capture systems. Fifteen healthy, non-pregnant women performed seven upright birthing positions. Hip, pelvis, and trunk kinematics were assessed using a 9-camera marker-based system and an 8-camera markerless system. Significant biomechanical differences were found between birthing positions. The “squat” position showed the most hip flexion and abduction, “B-Ball” had the greatest anterior pelvic tilt, and “all-fours” exhibited the most posterior tilt. For the trunk, “upright” led to the most extension, and “elbows bent knees” showed the most flexion. The markerless system aligned well with the marker-based system in the coronal and transverse planes but lacked sensitivity in the sagittal plane. This study highlights the biomechanical differences in upright birthing positions and emphasises the need for personalised birthing strategies. Understanding labour biomechanics is crucial for improving maternal and foetal well-being and reducing complications. By providing comprehensive and evidence-based information, women can make informed decisions about their birthing positions, enhancing outcomes and lowering the risk of maternal and neonatal complications globally.

## Introduction

Childbirth is a pivotal event in a woman’s life. Understanding the effects of birthing positions on the labour process and optimising these positions is crucial for improving maternal and foetal outcomes. ^1^ The World Health Organisation (WHO) encourages the adoption of upright positions (flexible sacrum positions) during labour due to their association with favourable childbirth outcomes. ^2^ However, the scientific understanding of the biomechanics of various birthing positions - recumbent, semi-recumbent, and upright - remains limited. ^3^ This limitation is primarily due to the constraints of methodologies available to collect and analyse biomechanical data and correlate them with maternal and neonatal outcomes. Methods such as pelvimetry, ^4,5^ magnetic resonance imaging, ^6,7^ and marker-based motion capture technology ^8,9^ have inherent limitations that prevent their use during labour. Additionally, these methods cannot effectively capture the dynamic and continuous nature of the birthing process. ^4–9^ This gap in knowledge impedes a comprehensive understanding of how different positions affect the birthing process. While computational modelling studies provide important insights, they cannot address this knowledge gap due to their inherent limitations, simplifications, and assumptions. ^10,11^

Several postural approaches have been introduced and marketed as alternatives for women’s positions during delivery. While these methods have shown enhanced women’s satisfaction through qualitative studies, ^12,13^ their effectiveness in promoting spontaneous vaginal births requires further investigation. ^3,14^ Theoretically, the optimal birthing position aligns the axis of foetal progression perpendicularly to the superior pelvic inlet, minimising obstacles by adjusting the lumbar dorsal hinge. ^8^ Different birthing positions exert unique biomechanical effects, influencing the progression of labour and maternal comfort. For instance, factors like pelvic tilt and sagittal plane hip angle significantly affect foetal descent and overall labour progression. ^3,8^

Research into the biomechanics of labour primarily concentrates on the angles of the pelvis and hips, ^10,11^ often neglecting equally crucial elements such as the spine, knees, and other vital factors, including body shape and weight. It is well understood that the human body functions in a domino-like fashion, where a modification in one area directly affects another in a kinetic chain. However, limited research has focused on the comprehensive biomechanical aspects of birthing positions, considering the global posture of the labouring woman ^9,15^ rather than just specific parts. Understanding this is vital for creating optimal conditions for foetal progression and improving childbirth outcomes. ^3^ The spine and pelvis are closely interconnected through the skeletal and muscular systems, and alterations in one area can influence the other. For instance, a change in the lumbar spine’s curvature can alter the tilt of the pelvis. ^16,17^ This is because the pelvis and the lower spine work together to support and balance the body, especially during movement and weight-bearing activities. Changes in spinal alignment, therefore, can lead to compensatory changes in the pelvic angle to maintain overall balance and function. ^18^ The hips, knees, ankles, feet, and lower back form an interconnected kinetic chain, crucial for movement and stability during childbirth. Dysfunction or limited motion in any part of this chain, such as the knees, can necessitate compensatory adjustments in adjacent areas, like the hips. For example, restricted knee mobility or angulations may alter the optimal hip positioning ^19^ and affect the choice of childbirth positions. Understanding these connections is essential in selecting positions that accommodate individual biomechanical needs and facilitate a smoother labour process. Among other factors, the shape and size of the pelvis, ^20^ as well as physical characteristics of lower extremities, which vary significantly between individuals, ^21^ play a crucial role in determining the effectiveness and comfort of various labour positions. Additionally, variations in abdominal weight and overall body mass result in shifts in the centre of gravity ^22,23^ and increased pressure on the spine and pelvis. ^24^ These biomechanical changes can make certain labour positions more or less favourable depending on the individual’s unique physical characteristics. Understanding these factors is essential for tailoring labour strategies to enhance comfort and facilitate the birthing process. These biomechanical variations highlight the necessity of tailoring and personalising positioning recommendations and support by maternal health providers to accommodate individual maternal needs and preferences. However, without the biomechanical knowledge to investigate these positions and understand their effects, due to the aforementioned limitations, we cannot provide biomechanically evidenced recommendations.

Recently, biomechanists around the world have started to investigate recumbent and semi-recumbent positions due to the practicalities in using motion capture techniques, and some upright position such as squatting. ^3,8,9,15^ To overcome the issues with marker-based systems that do not allow investigation in a real clinical environment, some attention has shifted towards newly developed markerless systems, ^8^ although there is not yet a comprehensive understanding of their capabilities.

In order to provide a better understanding of the available methods and their abilities to be used in real, outside-laboratory environments and investigate the most common upright birthing positions, this study compares two motion capture systems — reference standard marker-based and novel markerless systems—and evaluates their efficacy and potential benefits in assessing the biomechanics of the most commonly used globally flexible sacrum birthing positions. To the best of our knowledge, this is the first study to utilise markerless motion capture technology for this purpose, aiming to provide a deeper and more nuanced understanding of the biomechanics involved during different positions.

The study was guided by the following hypotheses and questions:

1. Hypothesis 1: Different upright birthing positions will exhibit distinctly different biomechanical characteristics of the spine, torso, pelvis, and hips.

Research Question 1: What are the specific biomechanics of upright birthing positions on the characteristics of the spine, torso, pelvis, and hips?

2. Hypothesis 2: The markerless motion capture system will provide comparable sensitivity to change among different upright birthing positions compared to the marker-based motion capture system.

Research Question 2: To what extent does the markerless motion capture system, compared to the marker-based, accurately capture and detect changes among different upright birthing positions?

## Material and Methods

### Design

A within-subjects, repeated measures design was used to analyse changes in postural kinematic parameters in seven upright birthing positions. The research was developed in collaboration with the public and stakeholders, ensuring it was conducted ‘with’ and ‘by’ them rather than merely ‘to,’ ‘about,’ or ‘for’ them. The stakeholders and the Patient and Public Involvement and Engagement (PPIE) group influenced the development of the study’s methodological approach and the choice of birthing positions to evaluate, focusing on those most widely used worldwide and the practicality of gathering data in a timely manner. By adopting this cooperative strategy, the study was made relevant and reflective of the needs and viewpoints of the intended audience. ^25,26^

### Participants

Healthy non-pregnant females (biological sex) of reproductive age ≥18 - ≤49 years old, ^27^ who were free from any injury, pain, illness, or medical condition that would limit their ability to perform the specific positions were recruited through campus-based advertisements and social media posts. In accordance with previously published rules of thumb for pilot studies, ^28–30^ a sample size of n=15 participants was determined for this exploratory study.

### Equipment

Data were collected synchronously using two motion capture systems: a 9-Oqus-camera marker-based motion capture system (Qualisys Medical AB, Sweden) and an 8-Micus-camera markerless motion capture system (Qualisys, Medical AB, Sweden). ^31^ Data from both systems were collected simultaneously in Qualisys Track Manager v2021.2 (Qualisys, Medical AB, Sweden), ensuring the systems were synchronised and calibrated in space and time by applying the same wand calibration. ^32^ Footage from the markerless system was recorded at 50Hz and data from the marker-based system was recorded at 100Hz, as per previous comparative studies. ^31–33^ Sampling frequencies vary due to the different capabilities of the two systems. Markerless systems typically use lower sampling frequencies due to the constraints of the frame rate of the RGB video cameras within the system. ^34^ Reducing the sampling frequency of the marker-based system to match the markerless system may affect the precision and accuracy of the marker-based data.^35^ A higher frame rate in the markerless system would increase the computational load during data processing, impacting real-time application.^34,35^

### Data Collection

Data was collected between 6^th^ July and 24^th^ October 2023. Participants attended a 45-minute data collection session in the University’s Motion Analysis Laboratory. Following participant consent, age, height, and weight were recorded. To maximise the quality of the data, all participants wore athletic style shorts and an adapted t-shirt which afforded access to the spine.

Data collection using the marker-based system requires the placement of passive retro-reflective markers using the calibrated anatomical system technique to afford tracking of segmental kinematics in 6 degrees of freedom. ^36^ Markers were attached using dermatological friendly double-sided adhesive tape, and were applied to the acromions, sternal notch, 7^th^ cervical vertebra (C7), anterior superior iliac spines (ASIS), posterior superior iliac spines (PSIS), greater trochanters, and the medial and lateral epicondyles. Clusters of four non-collinear markers were attached to the thighs and spinal clusters were applied level with the 7^th^ thoracic vertebra, 3^rd^ and 5^th^ Lumbar vertebra. ^37,38^ Using this configuration of markers, the study effectively delineated anatomical segments including the Lower Thoracic (LT), Upper Lumbar (UL) and Lower Lumbar (LL) spinal segments, a thorax (defined using the anatomical locations of the acromions, sternal notch, 7^th^ cervical vertebra and ASIS), a pelvis (defined using the anatomical locations of the ASIS and the PSIS), and hips (Fig 1).

**Figure 1.** Marker set used for the marker-based motion capture system. Note: In line with MedRxiv policy, the figures have been removed. Readers may contact the corresponding author to request access to these materials.

The seven upright birthing positions investigated were:

1. <u>Standing upright [*Upright*] (</u>Fig 2A): Participants were instructed to stand up straight with feet shoulder-width apart. They were asked to keep their palms facing their legs and look straight ahead at a specific point indicated to them, maintaining a natural and relaxed posture without stiffening their shoulders or arms.
2. <u>Leaning forward - support on the palms [*Palms*] (</u>Fig 2B): Participants were instructed to stand beside the bed and lean forward, placing their palms on the bed at a width that felt comfortable to them, with their elbows extended. They were asked to keep their knees straight and their feet at a width that was comfortable for them.
3. <u>Leaning forward - support on elbows [*Elbows, extended knees*] (</u>Fig 2C): As above, participants were instructed to stand beside the bed and lean forward, supporting their weight on their elbows and forearms, which were placed on the bed. They were asked to cross their palms, forming a triangle. The feet were positioned as described previously, with the knees kept straight.
4. <u>Leaning forward - support on elbows - knees bent [*Elbows, bent knees]* (</u>Fig 2D): Participants were instructed to stand beside the bed and lean forward, supporting their weight on their elbows and forearms, which were placed on the bed. They were asked to cross their palms, forming a triangle. Their feet were placed at a width that was comfortable for them, and they were to bend their knees to an angle of 90-120 degrees, according to their own comfort and preference.
5. <u>Deep squat [*Squat*] (</u>Fig 2E): Participants were instructed to stand to the side of the bed. With their palms placed on the bed for support, but not holding onto the bed, participants were asked to perform a deep squat within a comfortable range.
6. <u>Hands and knees [*All-4s*] (</u>Fig 2F): Participants were instructed to climb onto the bed and assume an all-fours position, with the spine in a neutral alignment and their palms placed directly under their shoulders, ensuring proper support and balance. Additionally, they were asked to look at a specific point to keep their neck straight, ensuring that the C7 marker and the sternal notch marker were not obscured. This alignment was crucial for accurate data collection and to maintain the integrity of the marker placements.
7. <u>Sitting on a birthing ball [*B-Ball*] (</u>Fig 2G): Participants were instructed to sit on a birthing ball with their legs spread wide to ensure balance and comfort. They were asked to place their hands on their thighs in a position that did not obstruct any markers. Additionally, they were directed to focus their gaze on a specific point that was indicated to them. The size of the ball was chosen according to the participant’s height. To maintain stability, participants ensured their feet were fully planted on the floor.

**Figure 2:** Birthing positions: 2A. Upright, 2B. Palms, 2C. Elbows with knees extended, 2D Elbows with knees bent, 2E. Squat, 2F. All-4s, 2G. B-Ball. Note: In line with MedRxiv policy, the figures have been removed. Readers may contact the corresponding author to request access to these materials.

During protocol development, and in combination with PPIE consultation, a formula was developed to set the height of the bed according to each participant’s height. This provided a practical solution, enabling standardisation relative to each participant’s stature, whilst also ensuring comfort and consistency during data collection. Maintaining a consistent reference point relative to body height is crucial for obtaining reliable kinematic data. The following formula was used:

*(Participant’s height in inches) + 1 = height of physio bed in cm*

Participants maintained each of the seven birthing positions for six seconds and assumed each position three times. If each position is considered as a static hold exercise, participants were required to remain motionless, counteracting the effects of gravity throughout the duration of the task. Taking into account the characteristics of participants who were ‘healthy’ but not necessarily athletically trained, and the number of required repetitions per task, six seconds was considered a suitable amount of time to ensure good data was collected without causing potential muscle fatigue amongst participants. To reduce bias and control for the potential influence of confounding factors, such as the effects of any extraneous variables that might affect the results on the outcome measures being studied, the order of the positions was randomised using the online Research Randomizer Tool (http://www.randomization.com).

There was a minimum of 30 second intervals between positions, with more offered if participants requested. Once participants had completed all seven birthing positions three times, the data collection session was complete.

### Data Processing and Analysis

The time domain of the measurements for both systems were time normalised to 101 data points. ^31^ As the marker-based system is considered the reference standard, the most stable two seconds of data was used for each trial, which was determined using manual event determination. Events for the markerless data were then synchronised with the events from the marker-based data.

#### Marker-Based Data

Anatomical frames were defined by landmarks positioned at the medial and lateral borders of the joint, from these right-handed segment co-ordinate systems were defined. The kinematics were calculated based on the Cardan sequence of XYZ equivalent to the joint coordinate system. ^39^ Raw kinematic data were exported from Qualisys Track Manager v2021.2 (Qualisys, AB, Sweden) to Visual3D (C-Motion Inc, USA) and filtered using a 6Hz Low Pass filter. Intersegmental spinal position (LT segment relative to UL segment, UL segment relative to LL segment and LL segment relative to the pelvis), trunk (trunk segment relative to the pelvis), pelvis (pelvis segment relative to the lab) and hip (thigh segment relative to the pelvis) angles were exported into Microsoft Excel, and the mean position for each trial were found.

#### Markerless Data

Theia3D (Theia Markerless Inc., Kingston, ON, Canada) is a deep learning algorithm-based approach to markerless motion capture that uses synchronised video data to perform 3D human pose estimation. ^40,41^ The raw video footage from 8 video cameras was processed using Theia3D (v2023.1.0.3161), from which 4×4 pose matrices of each body segment were exported for analysis into Visual3D (C-Motion Inc, USA). Theia3D embeds a multibody kinematic model consisting of two separate kinematic chains: one for the lower extremity and one for the upper extremity, and a separate head segment (https://www.theiamarkerless.ca/docs/model.html). For this study, from the upper extremity chain, the torso (trunk) segment was used, and from the lower extremity chain, the pelvis and hip joints were used. Trunk, pelvis and hip angles were exported from Visual3D (C-Motion Inc, USA) into Microsoft Office Excel 365 (Microsoft Corp, USA), and the mean position for each trial were found.

### Statistical Analysis

All statistical analyses were performed using SPSS v28 (IBM Corp., NY, USA). Descriptive statistics were used to describe the characteristics of participants.

To investigate the differences between birthing positions, one-way Repeated Measures Analysis of Variance (ANOVA) tests were performed for each motion capture system separately. This was also undertaken to determine the sensitivity of the markerless system i.e. whether statistically significant differences between birthing positions were the same. Mean joint angles (°), standard deviations (SD), main effects and effect sizes (partial eta squared, ηp^2^) were reported. Where a main effect was seen Least Significant Difference post-hoc pairwise comparisons were performed. Pairwise comparisons, mean differences (MD), and 95% confidence intervals of the mean differences (CI) were reported.

As a further step, to examine the sensitivity of the markerless system, and determine the extent to which data from the markerless motion capture systems matched that from the marker-based system, linear regression models were used for each outcome measurement. Due to be it being considered the reference standard, variables from the marker-based system were selected as dependent variables and those from the markerless system the independent/ predictor variables. The R square (R²) was reported as a goodness-of-fit measure (0.0 – 1.0); greater R² values represent smaller differences between the two systems. Significance values (p value) were also presented for each linear regression model. Statistical significance was set at the p<0.05 level throughout.

### Ethical Considerations and Data Protection

The study was approved by the University of Central Lancashire - Health Ethics Committee (HEALTH 01203). Data collection conformed to the Declaration of Helsinki, ^42^ and all information collected was kept strictly confidential, with participant anonymity maintained throughout. Data were collected, managed, analysed, and stored in accordance with General Data Protection Regulations (GDPR). Volunteers provided written informed consent prior to participation. During the consent process, participants were briefed on data protection measures and their rights concerning their data.

## Results

### Participant Characteristics

Fifteen healthy non-pregnant participants were included in the study; their characteristics are described in Table 1.

**Table 1:** Participant Characteristics.

| Characteristic | Mean (SD) | Range |
| --- | --- | --- |
| Age (years) | 31 (8.5) | 23 - 48 |
| Body mass (kg) | 69.5 (12.8) | 51.5 - 97 |
| Height (cm) | 166.0 (7.3) | 153.2 – 179.6 |
| History of pregnancy (n) | Yes n=7, No n=8 |  |
| History of giving birth (n) | Yes n=4, No n=11 |  |

### Differences Between Upright Birthing Positions: Markerless Motion Capture System

#### Hip

At the hip, significant main effects were observed in the sagittal, coronal and transverse planes (p<0.001; Table 2). In the sagittal plane, participants assumed the most amount of hip flexion in the “squat” position, and the most amount of hip extension in the “upright” position (Table 9). Apart from when comparing between “B-Ball” and “elbows, bent knees” (p=0.973), all other positions produced significantly different hip flexion-extension angles compared to each other (p<0.016; Table 3).

**Table 2:**
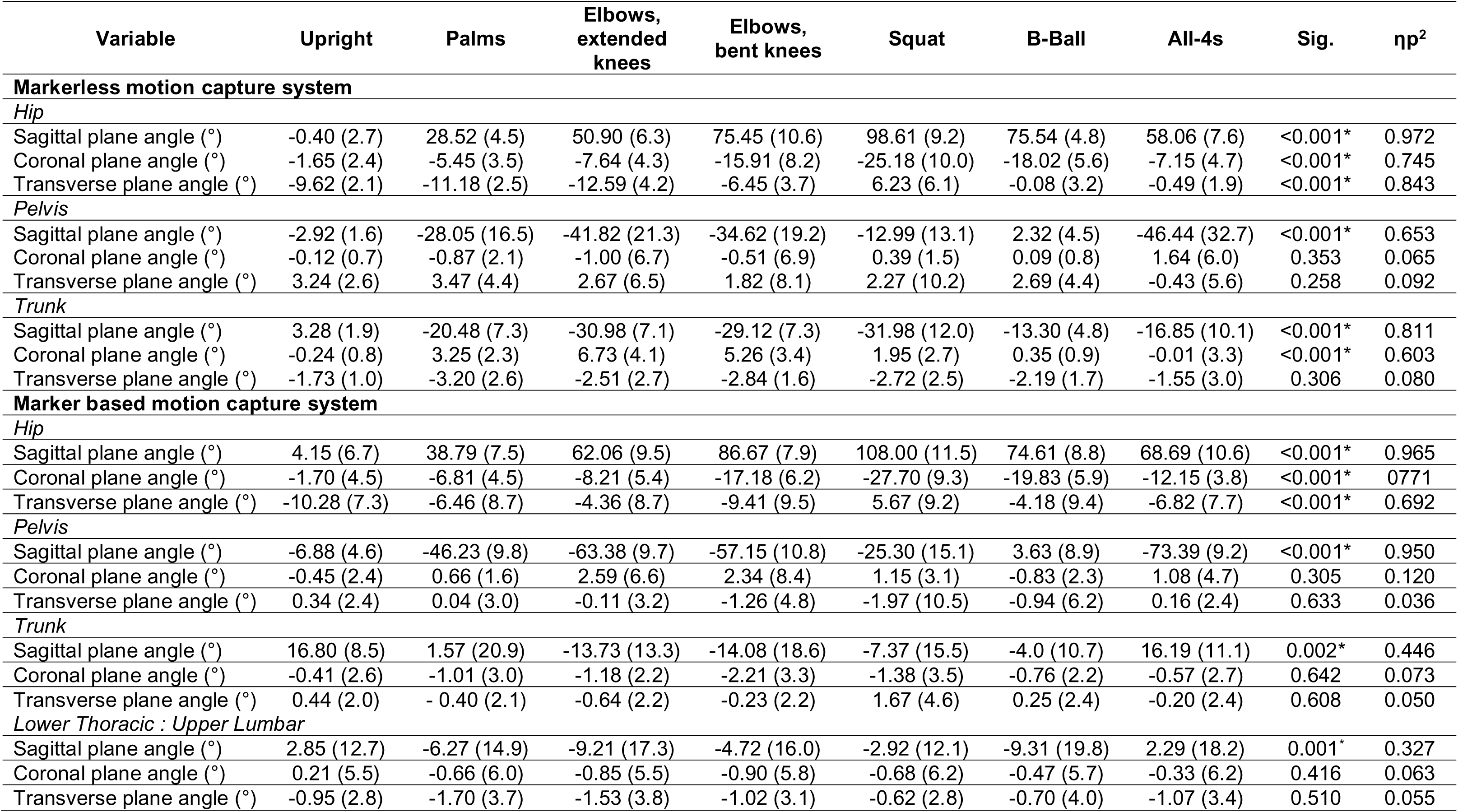
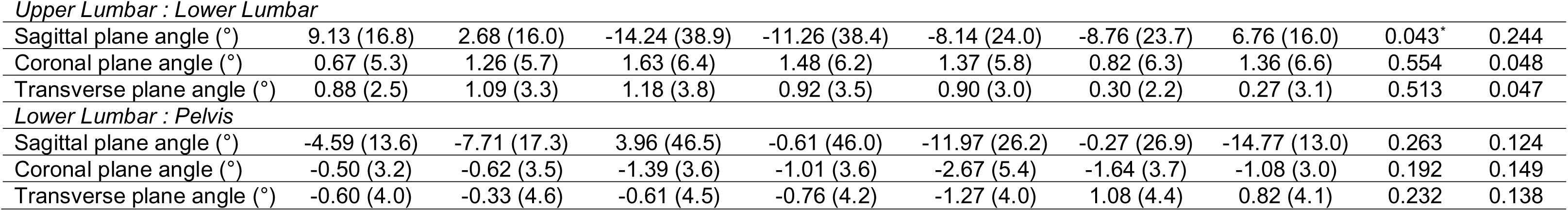
Mean (SD), main effects (Sig) and effect sizes (ηp^2^) for data from the markerless and marker-based motion capture systems. Significance level p<0.05. * Denotes significance.

| Variable | Upright | Palms | Elbows,<br>extended<br>knees | Elbows,<br>bent knees | Squat | B-Ball | All-4s | Sig. | $\eta^2$ |
| --- | --- | --- | --- | --- | --- | --- | --- | --- | --- |
| **Markerless motion capture system** |  |  |  |  |  |  |  |  |  |
| *Hip* |  |  |  |  |  |  |  |  |  |
| Sagittal plane angle (°) | -0.40 (2.7) | 28.52 (4.5) | 50.90 (6.3) | 75.45 (10.6) | 98.61 (9.2) | 75.54 (4.8) | 58.06 (7.6) | <0.001* | 0.972 |
| Coronal plane angle (°) | -1.65 (2.4) | -5.45 (3.5) | -7.64 (4.3) | -15.91 (8.2) | -25.18 (10.0) | -18.02 (5.6) | -7.15 (4.7) | <0.001* | 0.745 |
| Transverse plane angle (°) | -9.62 (2.1) | -11.18 (2.5) | -12.59 (4.2) | -6.45 (3.7) | 6.23 (6.1) | -0.08 (3.2) | -0.49 (1.9) | <0.001* | 0.843 |
| *Pelvis* |  |  |  |  |  |  |  |  |  |
| Sagittal plane angle (°) | -2.92 (1.6) | -28.05 (16.5) | -41.82 (21.3) | -34.62 (19.2) | -12.99 (13.1) | 2.32 (4.5) | -46.44 (32.7) | <0.001* | 0.653 |
| Coronal plane angle (°) | -0.12 (0.7) | -0.87 (2.1) | -1.00 (6.7) | -0.51 (6.9) | 0.39 (1.5) | 0.09 (0.8) | 1.64 (6.0) | 0.353 | 0.065 |
| Transverse plane angle (°) | 3.24 (2.6) | 3.47 (4.4) | 2.67 (6.5) | 1.82 (8.1) | 2.27 (10.2) | 2.69 (4.4) | -0.43 (5.6) | 0.258 | 0.092 |
| *Trunk* |  |  |  |  |  |  |  |  |  |
| Sagittal plane angle (°) | 3.28 (1.9) | -20.48 (7.3) | -30.98 (7.1) | -29.12 (7.3) | -31.98 (12.0) | -13.30 (4.8) | -16.85 (10.1) | <0.001* | 0.811 |
| Coronal plane angle (°) | -0.24 (0.8) | 3.25 (2.3) | 6.73 (4.1) | 5.26 (3.4) | 1.95 (2.7) | 0.35 (0.9) | -0.01 (3.3) | <0.001* | 0.603 |
| Transverse plane angle (°) | -1.73 (1.0) | -3.20 (2.6) | -2.51 (2.7) | -2.84 (1.6) | -2.72 (2.5) | -2.19 (1.7) | -1.55 (3.0) | 0.306 | 0.080 |
| **Marker based motion capture system** |  |  |  |  |  |  |  |  |  |
| *Hip* |  |  |  |  |  |  |  |  |  |
| Sagittal plane angle (°) | 4.15 (6.7) | 38.79 (7.5) | 62.06 (9.5) | 86.67 (7.9) | 108.00 (11.5) | 74.61 (8.8) | 68.69 (10.6) | <0.001* | 0.965 |
| Coronal plane angle (°) | -1.70 (4.5) | -6.81 (4.5) | -8.21 (5.4) | -17.18 (6.2) | -27.70 (9.3) | -19.83 (5.9) | -12.15 (3.8) | <0.001* | 0.771 |
| Transverse plane angle (°) | -10.28 (7.3) | -6.46 (8.7) | -4.36 (8.7) | -9.41 (9.5) | 5.67 (9.2) | -4.18 (9.4) | -6.82 (7.7) | <0.001* | 0.692 |
| *Pelvis* |  |  |  |  |  |  |  |  |  |
| Sagittal plane angle (°) | -6.88 (4.6) | -46.23 (9.8) | -63.38 (9.7) | -57.15 (10.8) | -25.30 (15.1) | 3.63 (8.9) | -73.39 (9.2) | <0.001* | 0.950 |
| Coronal plane angle (°) | -0.45 (2.4) | 0.66 (1.6) | 2.59 (6.6) | 2.34 (8.4) | 1.15 (3.1) | -0.83 (2.3) | 1.08 (4.7) | 0.305 | 0.120 |
| Transverse plane angle (°) | 0.34 (2.4) | 0.04 (3.0) | -0.11 (3.2) | -1.26 (4.8) | -1.97 (10.5) | -0.94 (6.2) | 0.16 (2.4) | 0.633 | 0.036 |
| *Trunk* |  |  |  |  |  |  |  |  |  |
| Sagittal plane angle (°) | 16.80 (8.5) | 1.57 (20.9) | -13.73 (13.3) | -14.08 (18.6) | -7.37 (15.5) | -4.0 (10.7) | 16.19 (11.1) | 0.002* | 0.446 |
| Coronal plane angle (°) | -0.41 (2.6) | -1.01 (3.0) | -1.18 (2.2) | -2.21 (3.3) | -1.38 (3.5) | -0.76 (2.2) | -0.57 (2.7) | 0.642 | 0.073 |
| Transverse plane angle (°) | 0.44 (2.0) | -0.40 (2.1) | -0.64 (2.2) | -0.23 (2.2) | 1.67 (4.6) | 0.25 (2.4) | -0.20 (2.4) | 0.608 | 0.050 |
| *Lower Thoracic : Upper Lumbar* |  |  |  |  |  |  |  |  |  |
| Sagittal plane angle (°) | 2.85 (12.7) | -6.27 (14.9) | -9.21 (17.3) | -4.72 (16.0) | -2.92 (12.1) | -9.31 (19.8) | 2.29 (18.2) | 0.001* | 0.327 |
| Coronal plane angle (°) | 0.21 (5.5) | -0.66 (6.0) | -0.85 (5.5) | -0.90 (5.8) | -0.68 (6.2) | -0.47 (5.7) | -0.33 (6.2) | 0.416 | 0.063 |
| Transverse plane angle (°) | -0.95 (2.8) | -1.70 (3.7) | -1.53 (3.8) | -1.02 (3.1) | -0.62 (2.8) | -0.70 (4.0) | -1.07 (3.4) | 0.510 | 0.055 |
| *Upper Lumbar : Lower Lumbar* |  |  |  |  |  |  |  |  |  |
| Sagittal plane angle (°) | 9.13 (16.8) | 2.68 (16.0) | -14.24 (38.9) | -11.26 (38.4) | -8.14 (24.0) | -8.76 (23.7) | 6.76 (16.0) | 0.043* | 0.244 |
| Coronal plane angle (°) | 0.67 (5.3) | 1.26 (5.7) | 1.63 (6.4) | 1.48 (6.2) | 1.37 (5.8) | 0.82 (6.3) | 1.36 (6.6) | 0.554 | 0.048 |
| Transverse plane angle (°) | 0.88 (2.5) | 1.09 (3.3) | 1.18 (3.8) | 0.92 (3.5) | 0.90 (3.0) | 0.30 (2.2) | 0.27 (3.1) | 0.513 | 0.047 |
| *Lower Lumbar : Pelvis* |  |  |  |  |  |  |  |  |  |
| Sagittal plane angle (°) | -4.59 (13.6) | -7.71 (17.3) | 3.96 (46.5) | -0.61 (46.0) | -11.97 (26.2) | -0.27 (26.9) | -14.77 (13.0) | 0.263 | 0.124 |
| Coronal plane angle (°) | -0.50 (3.2) | -0.62 (3.5) | -1.39 (3.6) | -1.01 (3.6) | -2.67 (5.4) | -1.64 (3.7) | -1.08 (3.0) | 0.192 | 0.149 |
| Transverse plane angle (°) | -0.60 (4.0) | -0.33 (4.6) | -0.61 (4.5) | -0.76 (4.2) | -1.27 (4.0) | 1.08 (4.4) | 0.82 (4.1) | 0.232 | 0.138 |

**Table 3:**
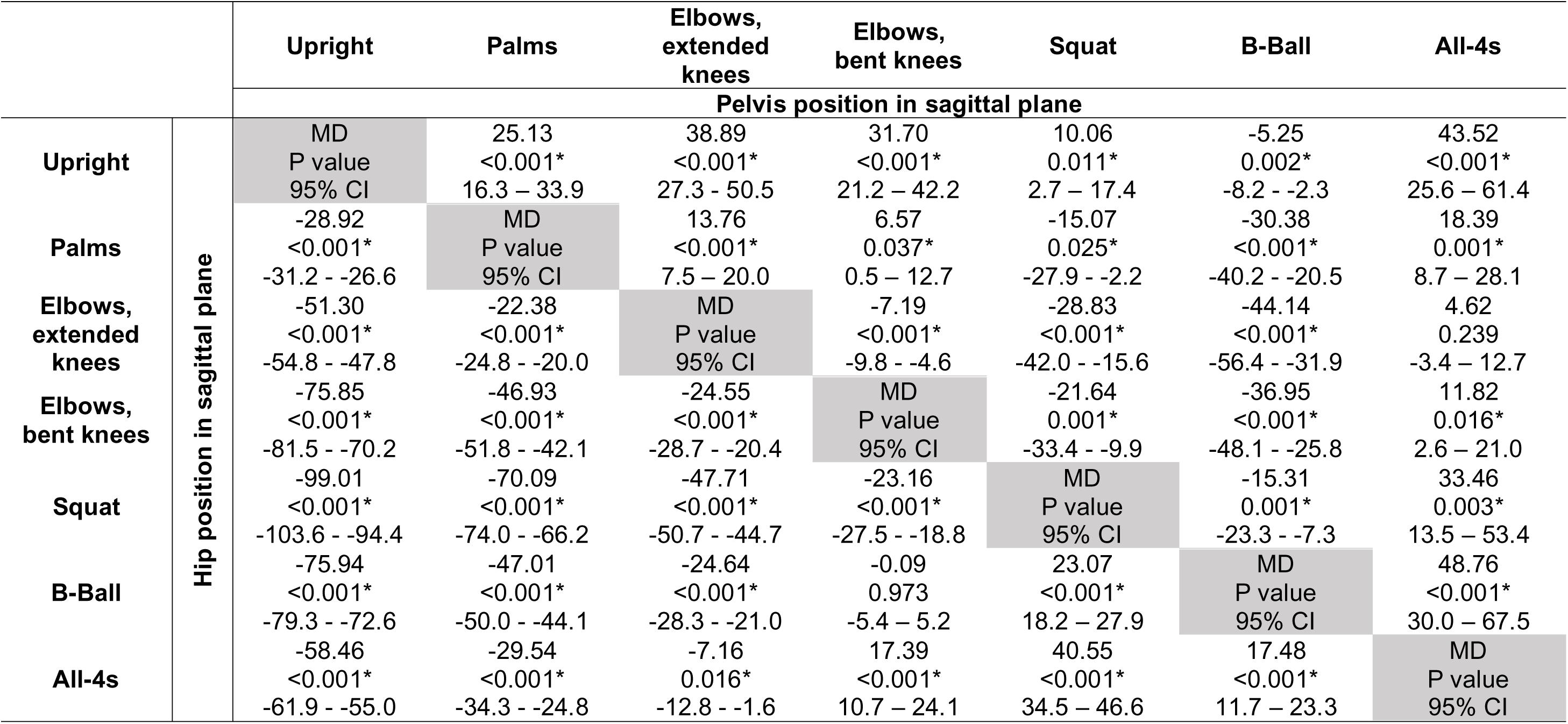
Markerless motion capture variables with main effects: Post-hoc pairwise comparisons (p value) of pelvis and hip position in the sagittal plane with mean differences (MD), and 95% confidence intervals (95% CI).

In the coronal plane, the “squat” position produced the greatest amount of hip abduction, while the “upright” position produced the least amount of hip abduction. Apart from when comparing between “all-4s” and “elbows, extended knees” (p=0.777), “all-4s” and “palms” (p=0.254), and “B-Ball” and “elbows, bent knees” (p=0.231; Table 4), all other positions were significantly different to each other (p<0.006).

**Table 4:**
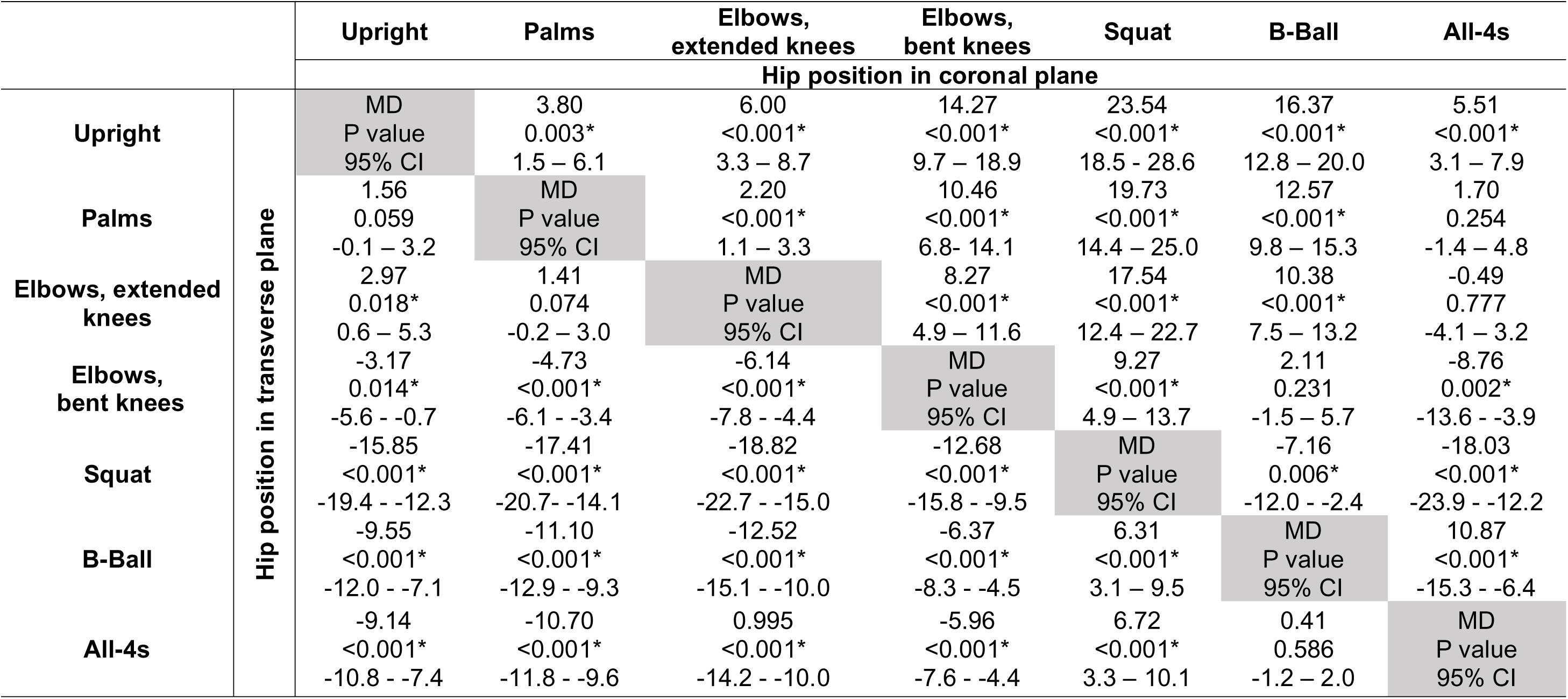
Markerless motion capture variables with main effects: Post-hoc pairwise comparisons (p value) of hip position in the coronal and transverse plane with mean differences (MD), and 95% confidence intervals (95% CI).

In the transverse plane, the “squat” position produced the greatest amount of external rotation while “elbows, extended knees” produced the greatest amount of internal rotation. Except when comparing between “upright” and “palms” (p=0.059), “palms” and “elbows, extended knees” (p=0.074), and “B-Ball” and “all-4s” (p=0.586; Table 4), all other positions were significantly different to each other (p<0.018).

#### Pelvis

At the pelvis, significant main effects were observed in the sagittal plane (p<0.001; Table 2). “B-Ball” produced the greatest amount of anterior tilt, while “all-4s” produced the greatest amount of posterior tilt. Except when comparing between “all-4s” and “elbows, extended knees” (p<0.239; Table 3), all other positions were significantly different to each other (p<0.037).

#### Trunk

For the trunk, significant main effects were observed in the sagittal and coronal planes (p<0.001; Table 2). In the sagittal plane, the “upright” position produced the greatest amount of trunk extension while the “squat” produced the most trunk flexion. All positions were significantly different to each other (p<0.002), except when comparing between “all-4s” and “palms” (p=0.055), “all-4s” and “B-Ball” (p=0.194), “elbows, bent knees” and “elbows, extended knees” (p=0.088), “squat” and “elbows, extended knees” (p=0.760), and “squat” and “elbows, bent knees” (p=0.317; Table 5).

**Table 5:**
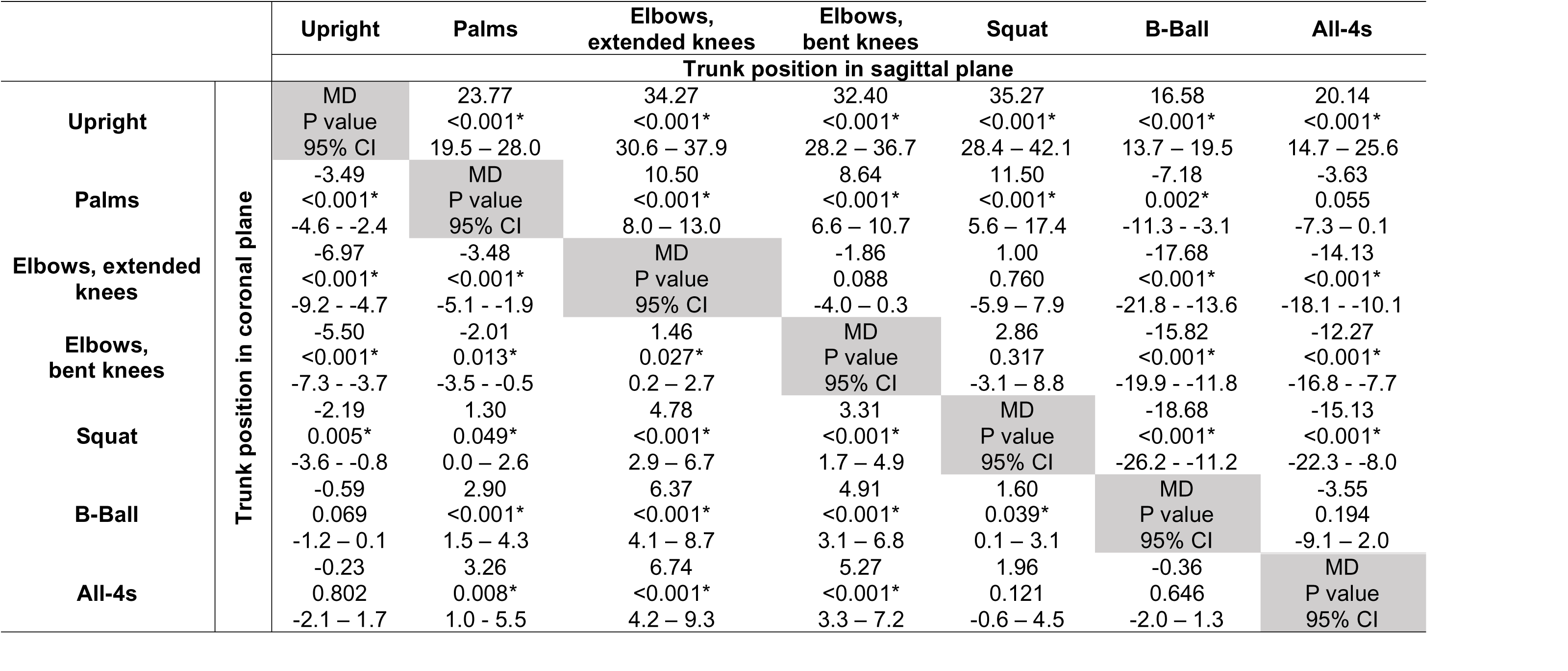
Markerless motion capture variables with main effects: Post-hoc pairwise comparisons (p value) of trunk position in the sagittal and coronal plane with mean differences (MD), and 95% confidence intervals (95% CI).

In the coronal plane, “all-4s” produced the least side flexion whilst “elbows, extended knees” produced the most side flexion. Except when comparing between “upright” and “B-Ball” (p=0.069), “upright” and “all-4s” (p=0.802), “all-4s” and “squat” (p=0.121), and “all-4s” and “B-Ball” (p=0.646; Table 5), all other positions were significantly different to each other (p<0.049).

### Differences Between Upright Birthing Positions: Marker-based Motion Capture System

#### Hip

At the hip, significant main effects were observed in the sagittal, coronal and transverse planes (p<0.001; Table 2) In the sagittal plane, the squat position produced the most hip flexion and the upright position produced the most hip extension. Except when comparing between “all-4s” and “elbows, extended knees” (p=0.207), and “all-4s” and “B-Ball” (p=0.173; Table 6), all other positions were significantly different from each other (p<0.007).

**Table 6:**
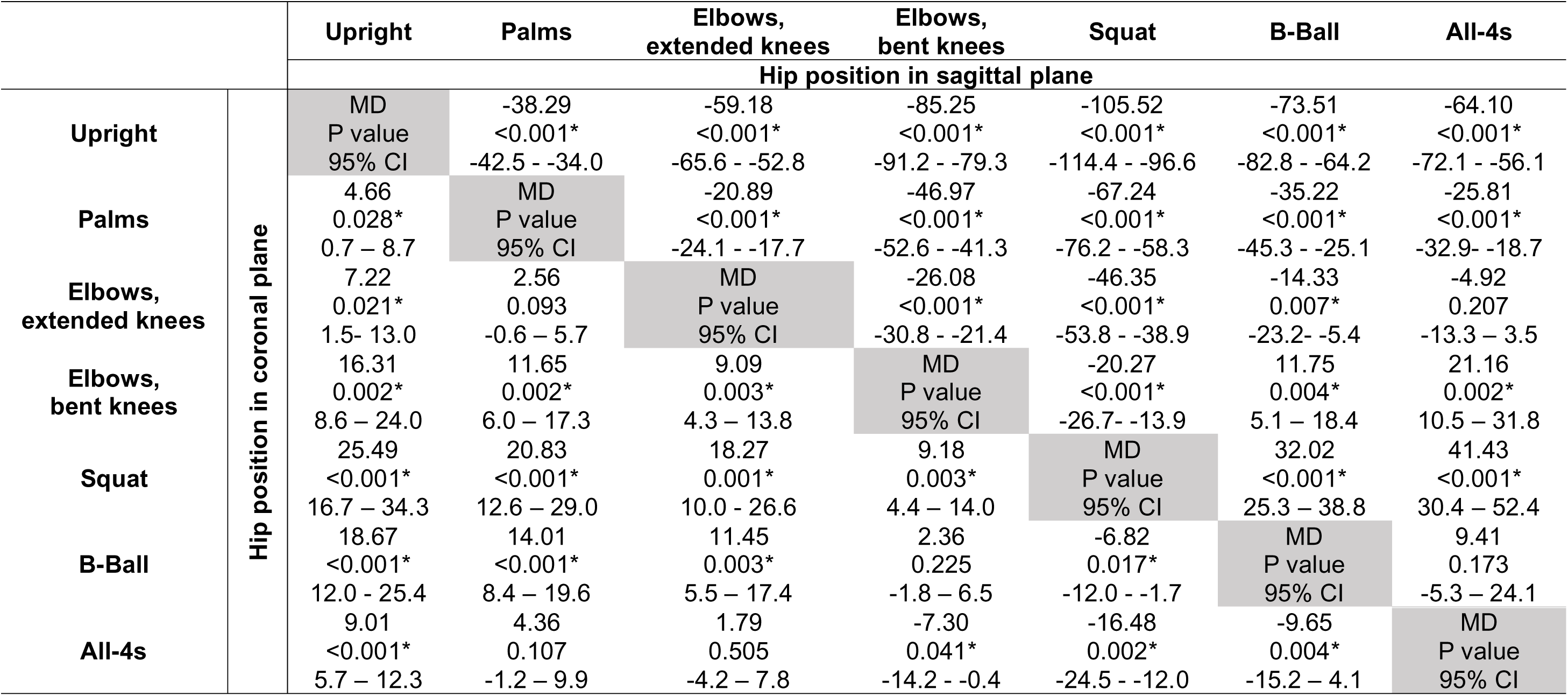
Marker based motion capture variables with main effects: Post-hoc pairwise comparisons (p value) of hip position in the sagittal and coronal plane with mean differences (MD), and 95% confidence intervals (95% CI).

In the coronal plane, the “squat” position produced the greatest amount of hip abduction, and the “upright” position produced the least amount. All positions were significantly different to each other (p<0.041), except when comparing between “all-4s” and “palms” (p=0.505), “all-4s” and “elbows, extended knees” (p=0.505), “B-Ball” and “elbows, bent knees” (p=0.225), and “palms” and “elbows, extended knees” (p=0.093; Table 6).

In the transverse plane the “squat” position produced the most external rotation and the “upright” position produced the most amount of internal rotation. Significant differences were observed when comparing the “upright” position with “palms” (p=0.018), “elbows, knees extended” (p=0.014), “squat” (p<0.001) and “B-Ball” (p=0.012; Table 7). “Palms” was also significantly different to “elbows, bent knees” (p=0.010) and “squat” (p=0.002). “Elbows, extended knees” was significantly different to “elbows, bent knees” (p=0.002) and “squat” (p=0.004). “Elbows, bent knees” was significantly different to the “squat” (p<0.001) and “B-Ball” (p=0.026). The “squat” position was significantly different to “B-Ball” (p=0.005) and “all-4s” (p<0.001).

**Table 7:**
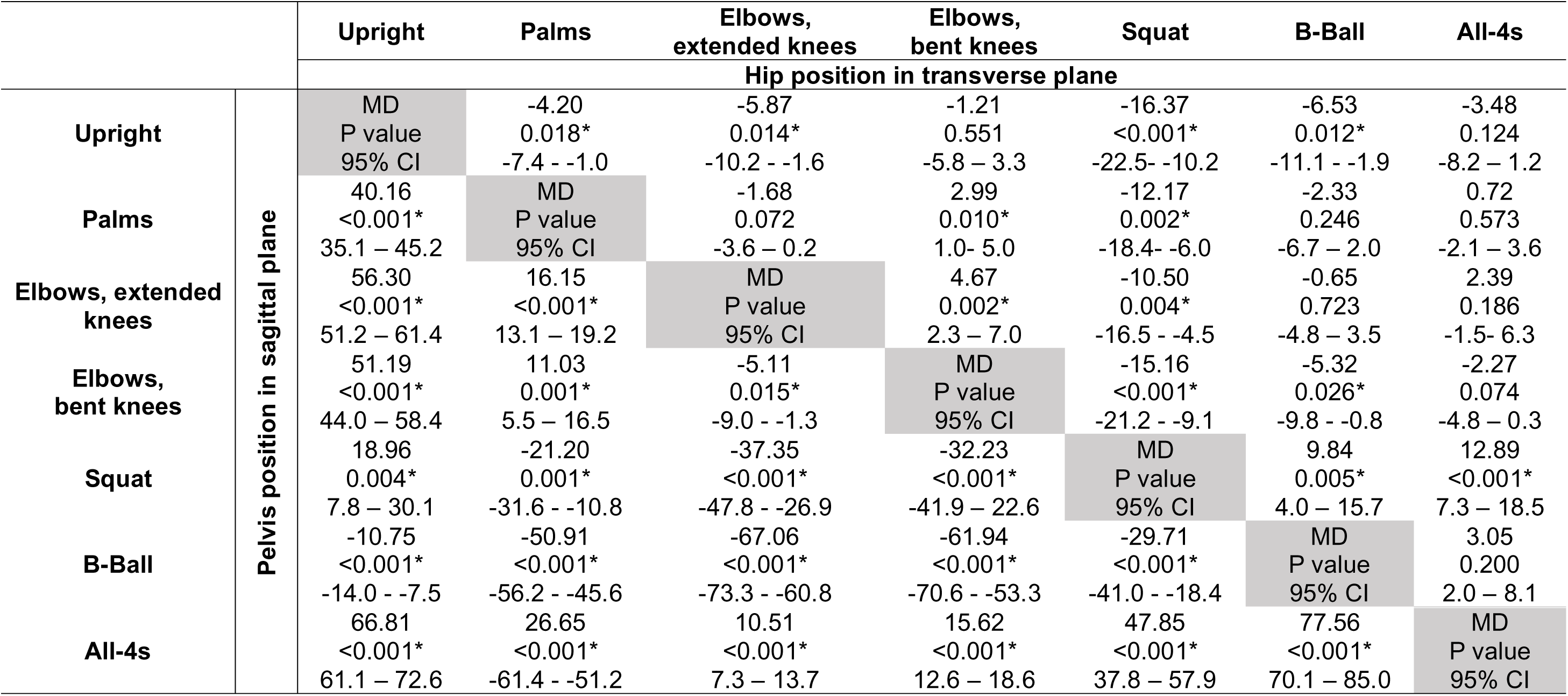
Marker based motion capture variables with main effects: Post-hoc pairwise comparisons (p value) of pelvis position in the sagittal plane and hip position in the transverse plane with mean differences (MD), and 95% confidence intervals (95% CI).95% CI).

#### Pelvis

At the pelvis, significant main effects were observed in the sagittal plane (p<0.001; Table 2). “B-Ball” produced the most anterior tilt and “all-4s” produced the most posterior tilt. All positions were significantly different to each other (p<0.015; Table 7).

#### Trunk

For the trunk, significant main effects were observed in the sagittal plane (p=0.002; Table 2). The “upright” position produced the most trunk extension and “elbows, bent knees” bent produced the most trunk flexion. The “upright” position was significantly different to all other positions (p<0.005) except “all-4s” (p=0.855; Table 8). “All-4s” was significantly different to “elbows, extended knees” (p<0.001), “elbows, bent knees” (p=0.005), “squat” (p=0.006) and “B-Ball” (p<0.001).

**Table 8:**
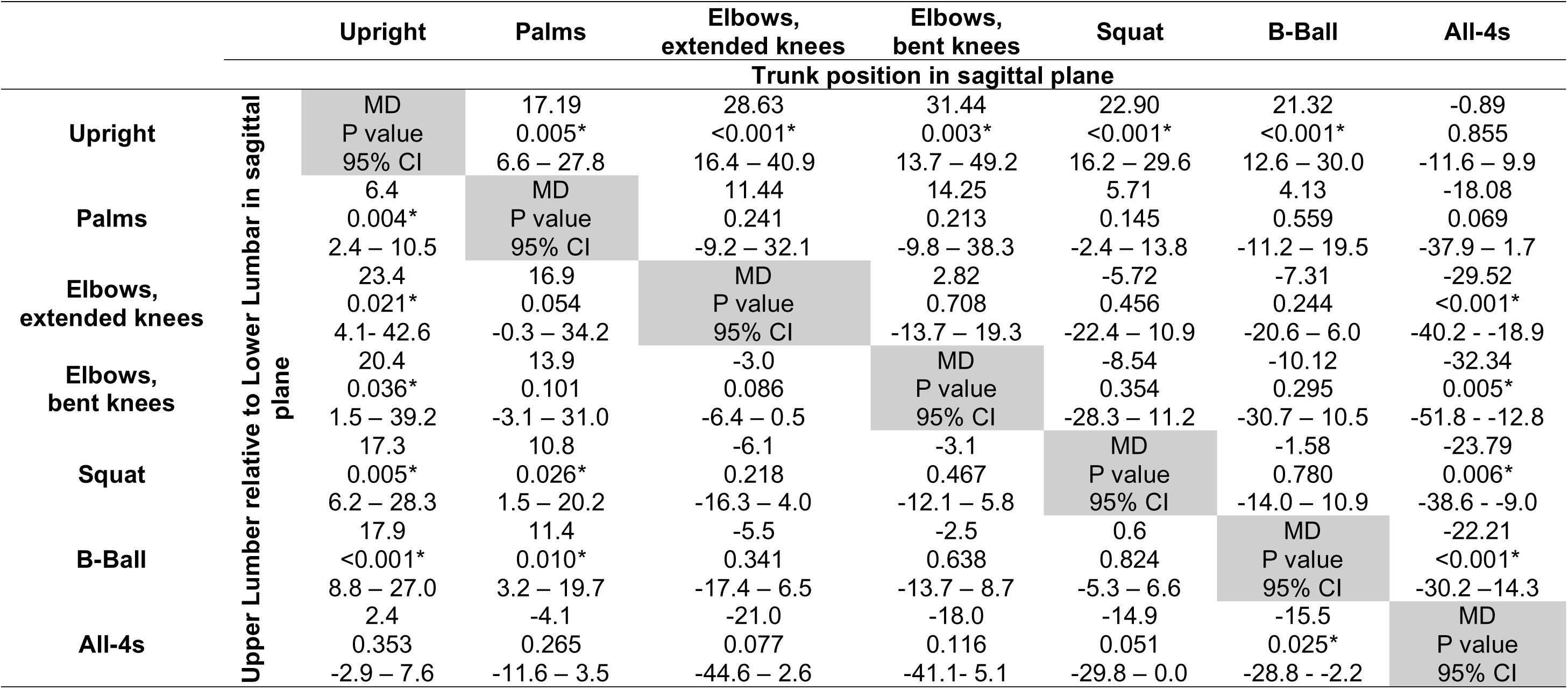
Marker based motion capture variables with main effects: Post-hoc pairwise comparisons (p value) of trunk position and Upper lumbar relative to lower lumbar in the sagittal plane with mean differences (MD), and 95% confidence intervals (95% CI). (95% CI).

#### Upper Lumbar segment relative to Lower Lumbar segment

Significant main effects were observed in the sagittal plane (p=0.043; Table 2). The “upright” position produced the most extension and the “elbows, extended knees” produced the most flexion. The “upright” position was significantly different to all other positions (p<0.036) except “all-4s” (p=0.353; Table 8). “Palms” was significantly different to “squat” and “B-Ball” (p<0.026), and “B-Ball” was significantly different to “all-4s”.

#### Lower Thoracic Segment relative to Upper Lumbar Segment

Significant main effects were observed in the sagittal plane (p<0.001; Table 2). The “upright” position was significantly different to all other positions (p<0.026) except “all-4s” (p=0.893; Table 10). “Palms” was significantly different to “all-4s” (p=0.010). “Elbows, extended knees” was significantly different to “elbows, bent knees”, “squat”, and “all-4s” (p<0.019; Table 9). “Elbows, bent knees” was significantly different to “all-4s” (p=0.012), and “B-Ball” was significantly different to “all-4s” (p=0.005). The “upright” position produced the most extension and the “B-Ball” position produced the most flexion (Table 10).

**Table 9::**
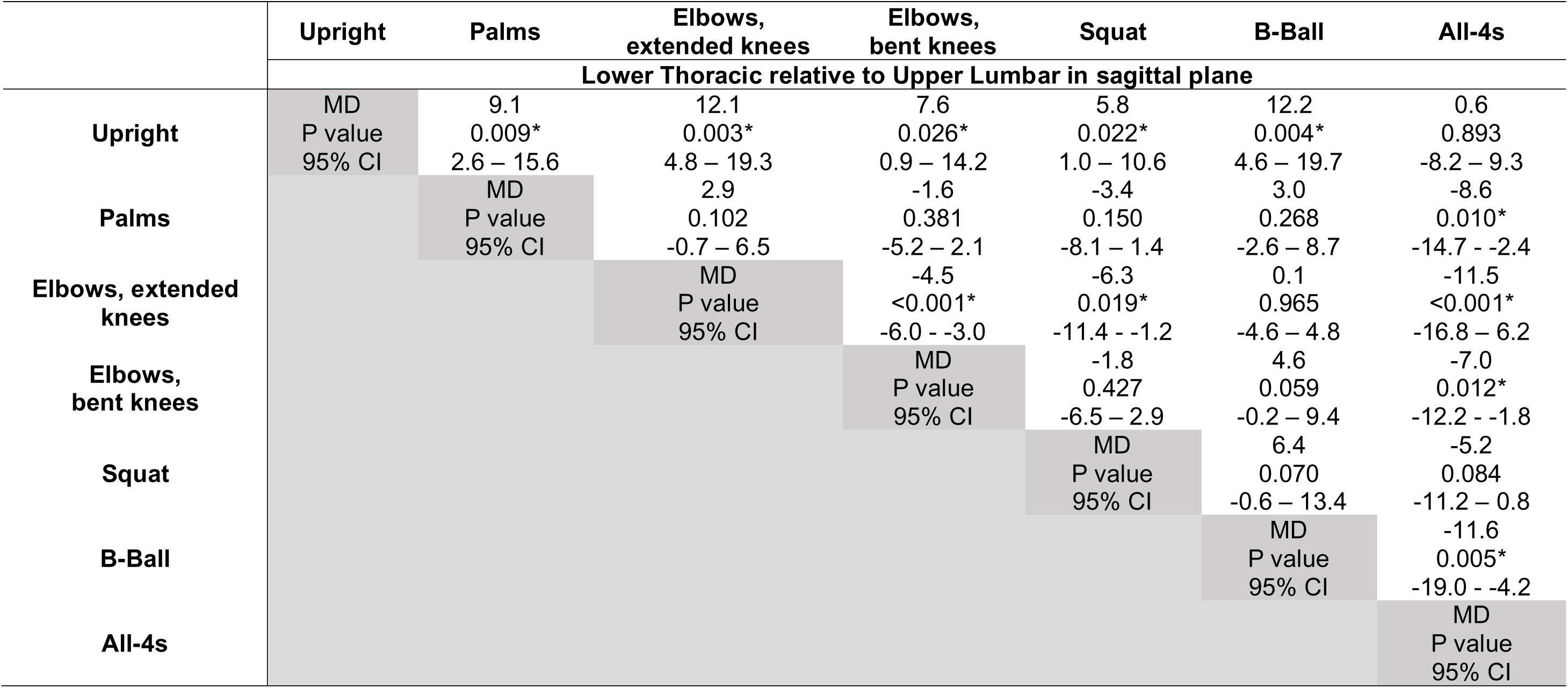
Marker based motion capture variables with main effects: Post-hoc pairwise comparisons (p value) of lower thoracic relative to upper lumbar in the sagittal plane with mean differences (MD), and 95% confidence intervals (95% CI).

**Table 10:**
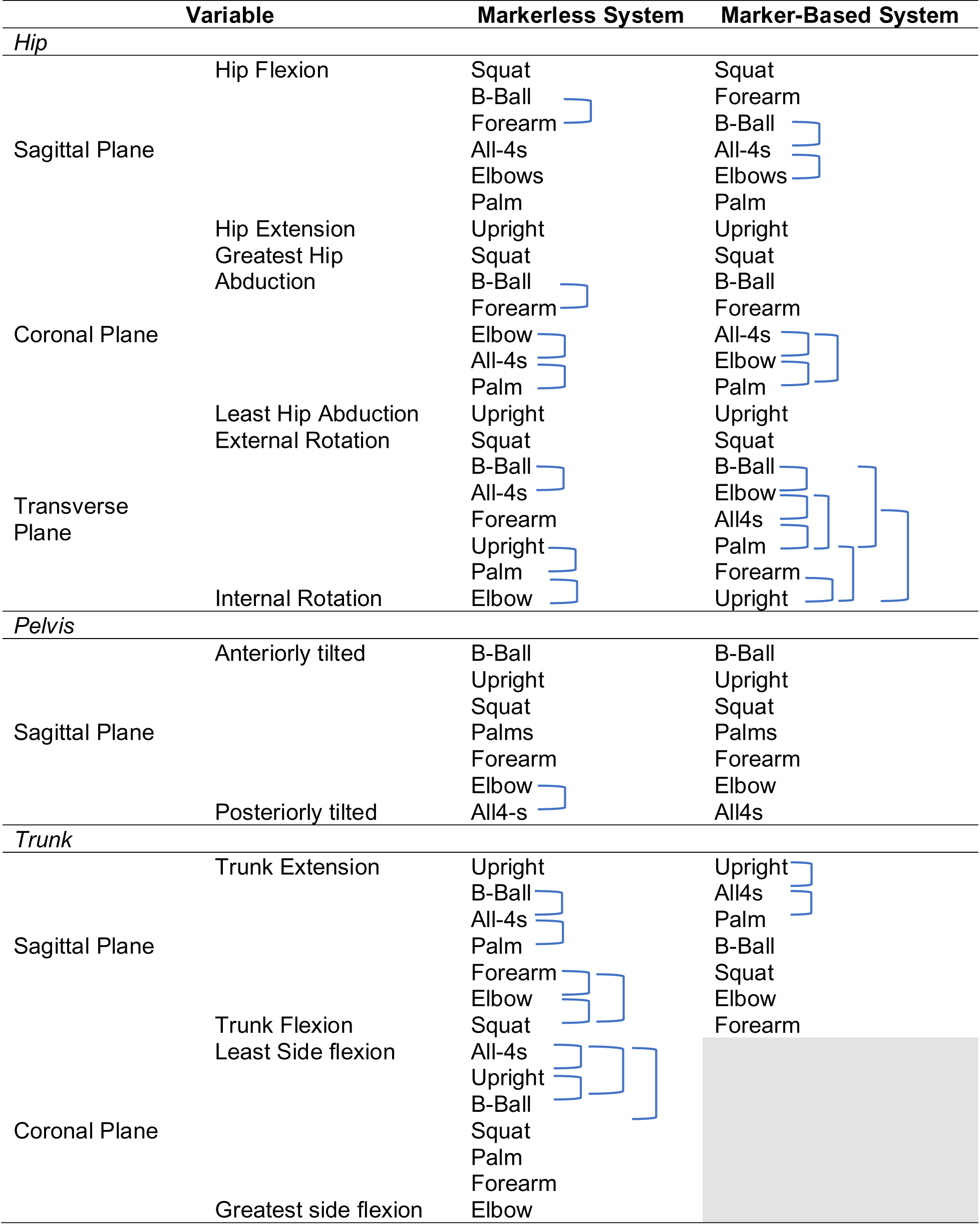
Comparative analysis of different positions across markerless and marker-based motion capture systems Closed bracket indicates no significant difference between the positions.

### Differences between Motion Capture Systems

#### Hip

The linear regression models revealed that the data from the markerless motion capture system matched the data from the marker-based system in only one out of seven positions for the hip joint variables in the sagittal plane. However, the markerless system performed better in the coronal plane, matching the data in six out of seven positions (Table 11). In the sagittal plane the markerless system corresponded with the marker-based system only in the “all-4s” position (R^2^ 0.575, p=0.002). In the coronal plane, the markerless system matched the marker-based system in the “upright” (R^2^ 0.799, p<0.001), “palms” (R^2^ 0.454, p<0.008), “elbows, extended knees” (R^2^ 0.639, p<0.001), elbows, bent knees” (R^2^ 0.714, p<0.001), “squat” (R^2^ 0.653, p<0.015), and “B-Ball” (R^2^ 0.630, p<0.001) positions, with a trend towards significance in “all-4s” (R^2^ 0.270, p<0.057).

**Table 11:** Linear regression analysis of angles by position in hip, pelvis and trunk regions.

|  | Upright |  | Palms |  | Elbows,<br>extended<br>knees |  | Elbows,<br>bent knees |  | Squat |  | B-Ball |  | All-4s |  |
| --- | --- | --- | --- | --- | --- | --- | --- | --- | --- | --- | --- | --- | --- | --- |
|  | R<sup>2</sup> | Sig. | R<sup>2</sup> | Sig. | R<sup>2</sup> | Sig. | R<sup>2</sup> | Sig. | R<sup>2</sup> | Sig. | R<sup>2</sup> | Sig. | R<sup>2</sup> | Sig. |
| *Hip* |  |  |  |  |  |  |  |  |  |  |  |  |  |  |
| Sagittal plane angle (°) | 0.133 | 0.199 | 0.007 | 0.773 | 0.266 | 0.071 | 0.226 | 0.119 | 0.439 | 0.073 | 0.228 | 0.099 | 0.575 | 0.002* |
| Coronal plane angle (°) | 0.799 | <0.001* | 0.454 | 0.008* | 0.639 | 0.001* | 0.714 | <0.001* | 0.653 | 0.015* | 0.630 | 0.001* | 0.270 | 0.057 |
| Transverse plane angle (°) | 0.025 | 0.588 | 0.001 | 0.915 | 0.027 | 0.591 | 0.010 | 0.759 | 0.171 | 0.309 | 0.018 | 0.661 | 0.059 | 0.404 |
| *Pelvis* |  |  |  |  |  |  |  |  |  |  |  |  |  |  |
| Sagittal plane angle (°) | 0.002 | 0.863 | 0.000 | 0.990 | 0.101 | 0.269 | 0.014 | 0.704 | 0.680 | 0.003* | 0.510 | 0.004* | 0.006 | 0.786 |
| Coronal plane angle (°) | 0.090 | 0.277 | 0.119 | 0.208 | 0.829 | <0.001* | 0.875 | <0.001* | 0.027 | 0.649 | 0.510 | 0.004* | 0.757 | <0.001* |
| Transverse plane angle (°) | 0.182 | 0.113 | 0.714 | <0.001* | 0.661 | <0.001* | 0.829 | <0.001* | 0.670 | 0.004* | 0.701 | <0.001* | 0.180 | 0.115 |
| *Trunk* |  |  |  |  |  |  |  |  |  |  |  |  |  |  |
| Sagittal plane angle (°) | 0.425 | 0.008* | 0.004 | 0.832 | 0.013 | 0.700 | 0.206 | 0.119 | 0.008 | 0.805 | 0.663 | <0.001* | 0.230 | 0.071 |
| Coronal plane angle (°) | 0.034 | 0.512 | 0.043 | 0.458 | 0.002 | 0.873 | 0.014 | 0.696 | 0.071 | 0.457 | 0.030 | 0.552 | 0.087 | 0.286 |
| Transverse plane angle (°) | 0.122 | 0.202 | 0.614 | <0.001* | 0.190 | 0.120 | 0.001 | 0.924 | 0.092 | 0.393 | 0.135 | 0.196 | 0.382 | 0.014* |

#### Pelvis

The linear regression models revealed that the data from the markerless motion capture system matched the data from the marker-based system in only two out of seven positions for pelvis variables in the sagittal plane. The markerless system performed better in the coronal and transverse planes, matching the data in four (coronal) and five (transverse) out of seven positions (Table 11). In the sagittal plane the markerless system corresponded with the marker-based system in the “squat” (R^2^ 0.680, p=0.003) and “B-Ball” positions (R^2^ 0.510, p=0.004). In the coronal plane, the markerless system agreed with the marker-based system in “elbows, extended knees” (R^2^ 0.829, p<0.001), elbows, bent knees” (R^2^ 0.875, p<0.001) and “B-Ball” (R^2^ 0.510, p=0.004) positions. In the transverse plane, the markerless system matched the marker-based system in “palms” (R^2^ 0.714, p<0.001), “elbows, extended knees” (R^2^ 0.661, p<0.001), “elbows, bent knees” (R^2^ 0.829, p<0.001), “squat” (R^2^ 0.670, p<0.004) and “B-Ball” (R^2^ 0.701, p<0.001) positions.

#### Trunk

The linear regression models revealed that the data from the markerless motion capture system matched the data from the marker-based system for variables in only two out of seven positions for trunk variables in the sagittal and transverse plane (Table 11). In the sagittal plane the markerless system corresponded with the marker-based system in the “upright” (R^2^ 0.425, p=0.008) and “B-Ball” positions (R^2^ 0.663, p<0.001). In the transverse plane, the markerless system matched the marker-based system in the “palms” (R^2^ 0.614, p<0.001) and “all-4s” (R^2^ 0.382, p=0.014) positions.

## Discussion

This study represents one of the first attempts to use both marker-based and markerless motion capture systems to assess the biomechanics of upright birthing positions. While the findings support our first hypothesis—that different upright birthing positions exhibit distinctly different biomechanical characteristics in the spine, torso, pelvis, and hips—they reject our second hypothesis. The markerless motion capture system did not provide comparable sensitivity to the marker-based system across all upright birthing positions.

### Markerless Motion Capture Sensitivity

While the markerless system showed promise in providing kinematic data in some positions, particularly in the coronal (hips and pelvis) and transverse (pelvis) planes, it did not consistently match the sensitivity of the marker-based system across all planes and positions. Although strong correlations were observed between the two systems in some positions, suggesting that markerless technology has the potential to be a viable alternative in some clinical applications, significant discrepancies were noted in the sagittal plane, particularly for the hip joint, where the markerless system did not perform as reliably. These discrepancies are more likely due to factors such as obstruction by body tissues, like the abdominal tissue or quadriceps, which can obscure the view of key joints in certain positions, as well as the inherent limitations in the system’s ability to accurately recognise joint positions in complex postures such as ‘all-4s’ and ‘elbows extended knees.’ Additionally, beyond potential issues with occlusion, limitations in algorithm accuracy ^43–45^ may further contribute to the system’s shortcomings. These findings, consistent with previous studies, ^43–45^ underscore the need for further refinement of the markerless system to ensure precision and reliability across all planes and positions. Despite its potential, the current limitations of markerless technology suggest that it may not yet be reliable enough for assessing the biomechanics of certain static positions, particularly for the joints and postures we analysed in this study.

### Biomechanical Variations Between Positions

Based on the above, we will discuss only the variations from the marker-based system. The results highlight that the biomechanics of each birthing position have unique effects on the hip in all planes, and in the sagittal plane for the pelvis, and trunk. Notably, the “squat” position exhibited the highest levels of hip flexion in the sagittal plane, hip abduction in the coronal plane, and external rotation in the transverse plane. This combination suggests that “squat” produces greater openness of the pelvic outlet, which could facilitate the descent of the foetus and aid in labour progression. However, a recent systematic review and meta-analysis does not show the squatting position during childbirth to be definitively beneficial, ^46^ The review indicated that, although the squatting position may decrease the need for instrumental delivery, it is also associated with an increased risk of caesarean section. ^46^ There was no clear evidence of a reduction in the duration of the second stage of labour or significant improvements in maternal or foetal outcomes. As there is no strong evidence either for or against squatting, women should be encouraged to choose the position they find most comfortable during labour. Furthermore, more studies are needed to better understand childbirth positions, especially in Western countries, as the majority of studies reviewed were conducted in Asia. ^46^

In contrast to squatting, the “upright” position produced the greatest hip extension in the sagittal plane and the least amount of hip abduction in the coronal plane, which may slightly reduce the dimensions of the pelvic outlet but provide a more comfortable and less physically demanding posture for some women. The “upright” position also promoted significant trunk extension in the sagittal plane, which increases the activation of the trunk extensor muscles to maintain stability by counteracting anterior loads. ^47^ Additionally, this posture encourages greater spinal extension, which could reduce discomfort for women who prefer an upright position. On the other hand, positions that induce greater trunk flexion, such as “elbows, bent knees,” may serve to redistribute weight and create a more forward-leaning posture, which could be advantageous during certain stages of labour.

The significant variations observed in pelvic tilt between positions are particularly important. The “B-Ball” position, which showed the greatest anterior pelvic tilt in the sagittal plane, could offer increased space in the pelvic inlet, facilitating the early stages of labour by allowing more room for the foetal head to engage. However, anterior pelvic tilt also increases lumbar lordosis (the inward curve of the lower back), which can strain the lower back, ^48,49^ potentially causing discomfort or misaligning the pelvis during labour. On the other hand, positions such as “all-fours” exhibited the greatest posterior pelvic tilt, which reduces the curvature of the lumbar spine, leading to a more neutral or slightly flexed lumbar position, which may help ease back pain and improve spinal alignment. ^49^ This posterior tilt could be particularly advantageous in later stages of labour when comfort and pain relief are priorities. These variations in pelvic tilt not only influence pelvic openness but also directly impact maternal comfort, especially in later stages of labour when comfort and pain relief are priorities. Positions like “all-fours,” which decrease lumbar lordosis, may be particularly beneficial for labour progression by creating more space in the birth canal and reducing lower back pain.

### Limitations of motion capture systems in labour settings

While both the marker-based and markerless motion capture systems provide valuable biomechanical data, significant limitations exist when considering their application in real-world labouring environments. The marker-based system, which requires the placement of retro-reflective markers on the participant’s body, poses several challenges for use in a clinical setting, particularly during labour. The presence of markers can be uncomfortable for a labouring woman, as they will disturb her from lying on her back or side or taking other positions, causing pressure, discomfort, and pain. Furthermore, markers can dislodge or fall off during movement, active positions, and handling by healthcare professionals or labour companions. In cases of interventions and/or emergencies and/or transfers, the time required to remove the markers would introduce unnecessary delays and complexities, which, apart from being impractical in critical situations, might also have a negative effect on safety. Additionally, the marker-based system relies on multiple cameras, cables, and processing units that would be challenging to set up in a labouring room and would add the risk of tripping hazards. Also, these individuals assist and touch the labouring woman, so their hands or bodies could inadvertently obscure the markers, making it impossible to gather reliable data in such a dynamic and cluttered environment. ^8,9,50–52^ The markerless motion capture system, although removes the need for physical markers, still faces similar practical limitations in a labouring room, including set up and unobstructed views. ^53,54^ Although not part of this study, it is worth mentioning that both motion capture systems face additional challenges when considering water births. For the marker-based system, the use of water would likely cause the markers to detach from the skin, rendering the system ineffective. For both systems, the presence of water would create an additional layer of complexity due to refraction and surface ripples, which could track the position of markers incorrectly and/or create substantial visual noise, affecting the system’s ability to generate reliable data. ^55,56^ Although it is possible to combine underwater cameras with a land-based camera system into a single motion capture setup, such an extremely sophisticated arrangement would further interfere with the environment, the practice, and the labouring woman’s comfort. ^57,58^

From the above, it is evident that no currently available motion capture system can effectively measure the biomechanics of labour due to the complexity of the equipment and methodologies, the dynamic nature of the birthing process, and the practical limitations posed by the labouring environment. Understanding the biomechanics of labour remains a critical need—not only because biomechanical complications are a major contributor to maternal and neonatal mortality, especially in low-income countries, ^10^ but also because it is unacceptable in 2024 that we still lack even a basic understanding of the mechanics involved in the beginning of human life. Addressing this knowledge gap is essential for improving childbirth outcomes globally.

### Study Limitations and Future Directions

As this was the first study assessing different birthing positions using two motion capture techniques, healthy non-pregnant women were selected as the study population. If pregnant women had been included, we would have needed to assess participants in the third trimester, as close as possible to their due date, due to the significant physiological ^59,60^ and anatomical changes that occur during pregnancy, particularly toward the latter stages of the third trimester. ^61,62^ This population would be considered vulnerable due to the potential physical exertion, ^63^ raising ethical concerns. Ensuring the safety of both the mother and foetus and taking into account the risk-benefit ratio is paramount. ^64^ Early-stage research using non-pregnant women allows for the exploration of biomechanical principles without exposing pregnant women to unnecessary risks. ^64^

Additionally, for an initial study providing essential groundwork, non-pregnant women offer a more controlled group for studying basic biomechanics. Their body weight, centre of gravity, and musculoskeletal function remain relatively stable compared to pregnant women, whose bodies undergo rapid changes. ^61,65^ This stability allows researchers to isolate and study the effects of specific birthing positions without the added variables introduced by pregnancy. This also helps establish baseline biomechanical data, testing the accuracy and sensitivity of motion capture systems before applying them to the more complex and variable biomechanics of pregnant women. Such a step is crucial for refining equipment and study methods.

Future studies should aim to replicate this analysis with pregnant participants to enhance ecological validity. This would account for changes in spinal curvature, particularly the lumbar curve, which increases by approximately 41% between 12 and 32 weeks of pregnancy. ^61^ Additionally, it would consider the influence of the hormone relaxin and pelvic mobility (however, experimental studies have not shown a direct relationship between high levels of relaxin and increased pelvic mobility or peripheral joint mobility in pregnant women), ^66^ and shifts in the centre of gravity. ^67^ In addition, it is crucial to understand the biomechanics of labour not only to enhance maternal and fetal well-being but also to reduce adverse outcomes and complications. ^68^ Researchers and clinicians should collaborate to create a method that allows for this analysis and knowledge, as current techniques show significant limitations for use during childbirth in clinical settings. ^52^ Understanding childbirth should be a top priority on the research agenda, not only because maternal and neonatal mortality remains unacceptably high, ^68–70^ but also because, from all the Sustainable Development Goals (SDG) for 2030, the SDG 3.1 target to reduce global maternal mortality is the only one that has failed. ^71^ Moreover, it is unacceptable to have such limited knowledge and understanding of the biomechanics of the beginning of human life.

## Conclusion

This study provides valuable insights into the biomechanics of various upright birthing positions, highlighting both the capabilities and limitations of current motion capture techniques. Future research should aim to refine these technologies and apply them to real-world labour environments. The findings demonstrate distinct biomechanical differences between positions, underscoring the need for personalised birthing strategies that take into account the unique biomechanical effects of each posture. Personalising support for women during childbirth is essential for their comfort, well-being, and the successful delivery of the baby. Maternal healthcare providers must have a solid understanding of anatomy and biomechanics, recognising how changes in one part of the body can affect others. For instance, variations in bone length, such as in the thighs, can significantly impact the mechanics of positions like squatting. A deep knowledge of these relationships is crucial for guiding women toward positions that not only support labour progress but also accommodate their unique body structures and preferences. Furthermore, it is important to recognise that different birthing positions may offer varying benefits depending on an individual’s characteristics, preferences, and the stage of labour. Women should receive comprehensive information and guidance during the antenatal period to make informed decisions about their birthing positions. Ultimately, these findings support the development of personalised, evidence-based birthing strategies that account for each woman’s biomechanical needs. By tailoring care to individual circumstances, we can improve maternal comfort, reduce complications, and enhance maternal and neonatal outcomes. A deeper understanding of childbirth biomechanics should remain a research priority, as it has the potential to significantly reduce maternal and neonatal mortality, a global challenge that persists today.

## Acknowledgements

This work was supported by the THRIVE Research Centre at UCLan. For open access, the author has applied a Creative Commons Attribution (CC BY) license to any Author Accepted Manuscript version arising. The authors would also like to thank Rebekah McCrimmon, Lecturer of Midwifery, for her clinical insights, and Dr. Jonathan Sinclair, Reader in Sport & Exercise Sciences and Nutrition & Exercise Sciences, for his invaluable contribution to the statistical analysis.

## Data availability statement

All data supporting the results of this study are available via the UCLanData repository DOI: 10.17030/uclan.data.00000475 Link: https://uclandata.uclan.ac.uk/475/

## Notes

### Competing Interest Statement

The authors have declared no competing interest.

### Author Declarations

The study was approved by the University of Central Lancashire - Health Ethics Committee (HEALTH 01203). Data collection conformed to the Declaration of Helsinki, 42 and all information collected was kept strictly confidential, with participant anonymity maintained throughout. Data were collected, managed, analysed, and stored in accordance with General Data Protection Regulations (GDPR). Volunteers provided written informed consent prior to participation. During the consent process, participants were briefed on data protection measures and their rights concerning their data.

